# Healthcare workers’ experiences of working on the frontline and views about support during COVID-19 and comparable pandemics: A rapid review and meta-synthesis

**DOI:** 10.1101/2020.06.21.20136705

**Authors:** Jo Billings, Brian Chi Fung Ching, Vasiliki Gkofa, Talya Greene, Michael Bloomfield

## Abstract

Healthcare workers across the world have risen to the demands of treating COVID-19 patients, potentially at significant cost to their own health and wellbeing. There has been increasing recognition of the potential mental health impact of COVID-19 on frontline healthcare workers and growing calls to provide psychosocial support for them. However, little attention has so far been paid to understanding the impact of working on a pandemic from healthcare workers’ own perspectives or what their views are about support. This rapid review identified 40 qualitative studies which have explored healthcare workers’ experiences and views from previous pandemics, including and comparable to COVID-19. Meta-synthesis of this qualitative data using thematic analysis derived eight key themes which transcended pandemics, time, and geographical boundaries. This pandemic is not unprecedented; the themes that arose from previous pandemics were remarkably resonant with what we are hearing about the impact of COVID-19 globally today. We have an opportunity to learn from the lessons of these previous pandemics, mitigate the negative mental health impact of COVID-19 and support the longer-term wellbeing of the healthcare workforce worldwide.

## Introduction

We have been hearing repeatedly from across the world about the mental health burden faced by frontline healthcare workers as they work to treat patients affected by COVID-19. Media representations have described frontline healthcare workers “on their knees” in response to the crisis, leading to forewarning of an ensuing mental health epidemic amongst the healthcare workforce (Ghebreyesus, 2020; Holmes et al., 2020).

COVID-19 has placed extreme demands on healthcare workers. They have been facing genuine threats to their own physical safety and indirectly to that of their families. They have had to manage higher numbers of patients with high mortality rates in a high-pressure environment. They have faced challenges in delivering care with strict infection control measures in place and not always with adequate personal protective equipment (PPE). Many have been redeployed into new roles or newly purposed wards so have been working in unfamiliar settings and without established social support from colleagues.

Healthcare workers are, for the most part, psychologically resilient professionals (Brooks et al., 2020); trained and experienced in dealing with illness and death. However, the mental health and psychological wellbeing of this group prior to the current COVID-19 pandemic was already being identified as a major healthcare issue, evidenced by the growing incidence of stress, burnout, depression, drug and alcohol dependence and suicide across all groups of health professionals, in many countries (Carrieri et al., 2018). High stress roles coupled with the unique demands of the current COVID-19 crisis have undoubtedly placed frontline healthcare workers at additional risk for mental health problems, with early reports from around the world indicating elevated rates of depression, anxiety, post-traumatic stress disorder (PTSD) and suicidality (Gunnell et al., 2020; Lai et al., 2020; Rossi et al., 2020).

In response to many military metaphors of healthcare workers “waging war on the front line” against COVID-19 there have been growing calls to mobilise mental health support for healthcare workers. However, there is currently a lack of evidence about what interventions are most helpful for staff working in such high risk occupational roles, with what evidence there is about effectiveness being mixed, and often demonstrating that well intentioned interventions, (i.e. debriefing) can actually increase the likelihood of developing mental health problems such as PTSD (Rose et al., 2002; National Institute for Health and Care Excellence, 2018).

Emerging literature from around the world has also demonstrated that staff may not prioritise psychological interventions in the peak phase of the crisis and may even be reluctant to engage with services offered to them (Chen et al., 2020). So, what is it like to work on the frontline and what support do frontline healthcare workers want during a pandemic such as COVID-19?

We set out to answer this question by providing a rapid review and meta-synthesis of qualitative literature describing healthcare workers’ experiences of working on the frontline and their views about psychosocial support, during Covid-19 or comparable pandemics, such as SARS, MERS or Ebola.

## Methods

We adhered to PRISMA (Moher et al., 2009) guidance throughout this review.

### Search strategy and selection criteria

We identified eligible papers through searches on Medline, PsychINFO and PubMed. Final searches took place on May 5, 2020. Key search terms were related to the population (i.e. healthcare workers), the intervention (i.e. psychosocial, psychological or emotional and experiences, support, intervention or help) and a number of disease specific key terms (i.e. COVID-19, SARS, MERS, H1N1, Ebola). (See appendix for our full list of search terms). We hand searched reference lists of included papers and grey literature to identify other potentially relevant studies.

We included papers which reported original, published, qualitative research describing frontline healthcare workers’ experiences of working during a pandemic, and/or views of psychosocial support offered to them. This included mixed-methods studies where data on the qualitative component of the study was available. We excluded studies where less than 50% of the participants were frontline healthcare workers and we could not extract data for healthcare workers alone. Due to the rapidity of this review, only papers published in English were included, although retrieved papers covered a global context.

### Data screening and extraction

We removed duplicate articles then titles and abstracts of retrieved papers were screened for relevance by two independent reviewers. The full texts of remaining articles were then read by the two independent reviewers. We resolved any discrepancies about papers to be included at any stage through discussion between the two reviewers and the first author. Data from the selected papers was extracted onto a pre-designed data extraction template by the two reviewers (see Table 1). We included all papers in the qualitative meta-synthesis.

**Table 1.** Selected studies.

| Author (year) | Country | Virus type | Sample | Study design (method of analysis) |
| --- | --- | --- | --- | --- |
| Aghaizu et al. (2011) | England, Hungary, Germany and Greece | SARS-CoV | 49 healthcare workers | 6 focus groups (thematic analysis) |
| Al Knawy et al. (2019) | Saudi Arabia | MERS-CoV | 28 mixed healthcare workers (9 management decision-makers and 19 healthcare workers) | Individual interviews and focus groups (process evaluation and thematic analysis) |
| Andertun et al. (2016) | Sierra Leone | Ebola | 9 (8 nurses and 1 physician) | Individual narrative and focus group interviews (qualitative content analysis) |
| Bensimon et al. (2007) | Canada | SARS-CoV | 67 mixed healthcare workers (25 healthcare providers - paramedics, respiratory therapists, social workers, physicians, nurses) | Semi-structured interviews (grounded theory although not explicitly stated) |
| Bergeron et al. (2006) | Canada | SARS-CoV | 941 community nurses | Open-ended questionnaire (thematic analysis) |
| Broom et al. (2017) | Australia | Ebola | 21 (8 consultants and 13 nurses) | Semi-structured interviews (thematic analysis) |
| Chen et al. (2020) | China | COVID-19 | 13 medical staff | Interview surveys (not stated) |
| Chiang, et al. (2007) | Taiwan | SARS-CoV | 21 nurses | Focus groups (thematic analysis) |
| Chung, et al. (2005) | Hong Kong | SARS-CoV | 8 nurses | 'Focused but non-structured talking technique' (Colaizzi's (1978) phenomenological method) |
| Erland & Dahl (2017) | Sierra Leone | Ebola | 11 midwives | Semi-structured interviews (thematic cross-case analysis) |
| Gershon et al. (2016) | West Africa | Ebola | 16 health care volunteers | Semi-structured interviews (thematic analysis) |
| Guimard et al. (1999) | Democratic Republic of Congo | Ebola | 27 nurses | Focus groups (not stated) |
| Hewlett & Hewlett (2005) | Uganda and Republic of Congo | Ebola | Mixed healthcare workers (Uganda - 6 individual nurses and 2 focus groups of nurses & healthcare workers) (Congo - 4 individual nurses and 2 focus groups of nurses & healthcare workers) | Open-ended and semi-structured interviews and focus groups (not stated) |
| Im et al. (2018) | South Korea | MERS-CoV | 8 nurses | Interviews (not stated) |
| Ives et al. (2009) | UK | General influenza pandemic | 64 healthcare workers | Focus groups and interviews (thematic analysis although not explicitly stated) |
| Kim (2018) | South Korea | MERS-CoV | 12 nurses | In-depth interviews (Colaizzi's (1978) phenomenological method) |
| Koh et al. (2012) | Singapore | SARS-CoV/H1N1 | 10 nurses | Semi-structured interviews (thematic analysis) |
| Lam & Hung (2012) | Hong Kong | Swine Flu | 10 nurses | Semi-structured interviews (content analysis) |
| Lamb (2018) | West Africa | Ebola | 14 mixed healthcare workers (doctors, nurses, allied health professionals and support personnel (health care assistants, biomedical scientists, PPE monitors and drivers)) | Semi-structured interviews (grounded theory) |
| Lau & Chen (2004) | Hong Kong | SARS-CoV | 1 (nurse manager) | Structured reflection (not stated) |
| Lee et al. (2005) | Taiwan | SARS-CoV | 26 nurses | Focus groups, semi-structured interviews and a questionnaire (not stated) |
| Liu & Liehr (2009) | China | SARS-CoV | 6 nurses | Descriptive exploratory qualitative study (content analysis) |
| Locsin et al. (2009) | Uganda | Ebola | 15 nurses | Written narratives (van Manen's (1990) hermeneutic phenomenological analysis) |
| McMahon et al. (2017) | Sierra Leone | Ebola | Health Volunteers (13 focus group discussions – exact number of participants not stated) | Focus groups (adapted framework approach) |
| Mok et al. (2005) | Hong Kong | SARS-CoV | 10 nurses | Interviews (content analysis) |
| Moore et al. (2005) | Canada | SARS-CoV | 105 mixed healthcare workers (occupational health staff, infection control practitioners, physicians, clinical nursing staff, allied health professionals (e.g. respiratory therapists, laboratory technicians, physiotherapists), support staff, hospital managers) | Focus groups (not stated) |
| O'Sullivan et al. (2009) | Canada | SARS-CoV | 100 nurses | Focus groups (thematic analysis) |
| Pearce et al. (2011) | Australia | General influenza pandemic | 19 (9 nurses, 10 GPs) | Interviews & focus groups (not stated) |
| Raven et al. (2018a) | Sierra Leone | Ebola | 25 frontline healthcare workers | In depth interviews, semi-structured interviews and observation (framework analysis) |
| Raven et al. (2018b) | Sierra Leone | Ebola | 25 mixed healthcare workers | In-depth interviews (framework analysis) |
| Rubin et al. (2016) | West Africa | Ebola | 51 mixed healthcare workers (30 Public Health England staff, 21 non-governmental organization (NGO) staff) | Telephone interviews (thematic analysis although not explicitly stated) |
| Sarikaya & Erbaydar (2007) | Turkey | Avian Flu | 17 mixed healthcare workers (12 doctors, 3 allied health personnel, 1 midwife, 1 nurse). | Interviews (thematic analysis) |
| Shaw et al. (2006) | Australia | General influenza pandemic | 60 GPs | Semi-structured interviews (thematic analysis although not explicitly stated) |
| Shih et al. (2007) | Taiwan | SARS-CoV | 200 Nurses | Semi structured interviews and open-ended questionnaire (thematic analysis) |
| Shih et al. (2009) | Taiwan | SARS-CoV | 70 nurse leaders | Focus group interviews (content analysis) |
| Smith et al. (2017) | USA | Ebola | 37 (any staff member who participated in the care of the EVD patients who were treated at the NBU during 2014.) | Semi-structured interviews (not stated) |
| Taylor et al. (2018) | USA | General influenza pandemic | 46 local health department staff (aimed for half the focus groups to be with frontline local health department staff) | Focus groups (thematic analysis) |
| von Strauss et al. (2017) | West Africa | Ebola | 44 nurses | Open-ended questionnaire (not stated) |
| Wong et al. (2011) | Hong Kong | H1N1 | 10 mixed healthcare workers | Semi-structured interviews (thematic analysis) |
| Yin & Zeng (2020) | China | COVID-19 | 10 nurses | Semi structured interviews (category analysis) |

### Quality appraisal

We assessed the quality of the studies included in the meta-synthesis using the Critical Appraisal Skills Programme (CASP; 2017) qualitative research checklist (see Table 2). The quality appraisal was carried out by two independent reviewers and discrepancies were resolved through discussion. The CASP checklists are designed to be used as educational pedagogic tools and therefore are not intended to derive a quantitative rating for quality. In this review, we have followed CASP guidance and the methods described by Lachal et al. (2017) to describe whether studies met, partially met, or did not meet the CASP criteria. This information is provided to enable the reader to judge study quality for themselves (see appendix). Of note, these ratings reflect what is included in the available report of the study and may not necessarily reflect detail that was attended to in the research process but not necessarily written up in the presented paper.

**Table 2.**
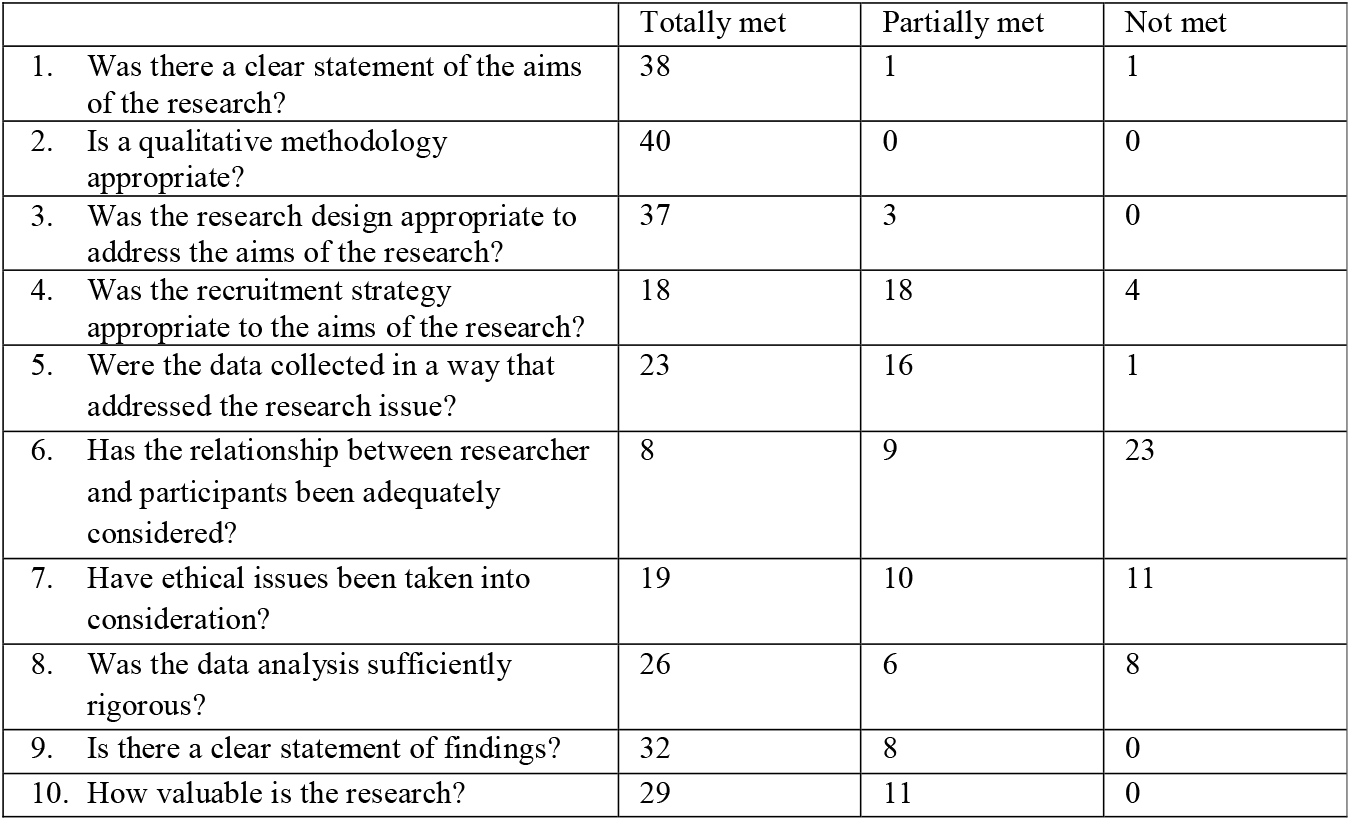
Quality of selected studies – number of studies meeting each CASP criteria.

We did not exclude any studies from the meta-synthesis based on their quality. This is a new and emerging topic of concern and we did not want to exclude smaller scale studies from less well-established settings given the insight they could potentially offer to this review question.

### Meta-synthesis

We followed guidance provided by Lachal et al. (2017) on synthesising qualitative literature in psychiatry. We extracted data from the results sections of papers (or general text in the case of published commentaries or reflective accounts) where information was given about healthcare workers’ experiences and/or views of any form of psychosocial support provided during their work in response to a pandemic. This data was exported into NVivo Pro version 12 and analysed thematically (Braun & Clarke, 2006).

In keeping with traditional thematic analysis, we sought initial immersion in the data by reading and re-reading all the papers. We developed an initial coding frame from ten of the most immediately relevant and current papers. The coding frame was further developed and refined through coding of the full 40 papers, looking for shared themes, but also nuances and exceptions within the themes. Adhering to the principles of inductive methodology, we sought to derive our themes from the data, in this case the themes and examples given in the original papers, but then to synthesise these findings and develop an overarching set of themes and sub-themes which captured the experiences and views of frontline healthcare workers across the studies.

### Reflexivity

Reflexivity is important in all qualitative research and enables the reader to consider the validity of any qualitative analysis by better understanding the composition and position of the research team who have produced it. This research team is made up of a diverse group, representing different clinical specialities, career stages and cultural backgrounds. JB is a Consultant Clinical Psychologist and Associate Clinical Professor, specialising in trauma, PTSD and the mental health and wellbeing of high-risk occupational groups. JB is also a specialist qualitative researcher with extensive experience of conducting qualitative research, systematic reviews and meta-syntheses. BCFC and VG are both MSc students in Clinical Mental Health. Both have received extensive training on conducting systematic reviews. TG is a Senior Lecturer specialising in PTSD and responses to mass traumatic events. MB is a UCL Excellence Principal Clinical Research Fellow and Honorary Consultant Psychiatrist at the Traumatic Stress Clinic, a specialist service for post-traumatic stress disorder in London, and University College London Hospitals NHS Foundation Trust. As such we brought a mix of different perspectives and experience to this topic.

## Results

A total of 896 records were initially returned. 34 articles were identified through other sources (hand searching of reference lists). After de-duplication, the titles and abstracts of 700 articles were screened by the two reviewers. Of these, 604 were agreed to not be relevant, resulting in 96 studies which were read in full by the two reviewers. At this stage, 56 studies were excluded as either the wrong study design (n=56) or not having relevant outcomes (n=11). This resulted in 40 papers which we included in the review and meta-synthesis (see figure 1).

**Figure 1.**
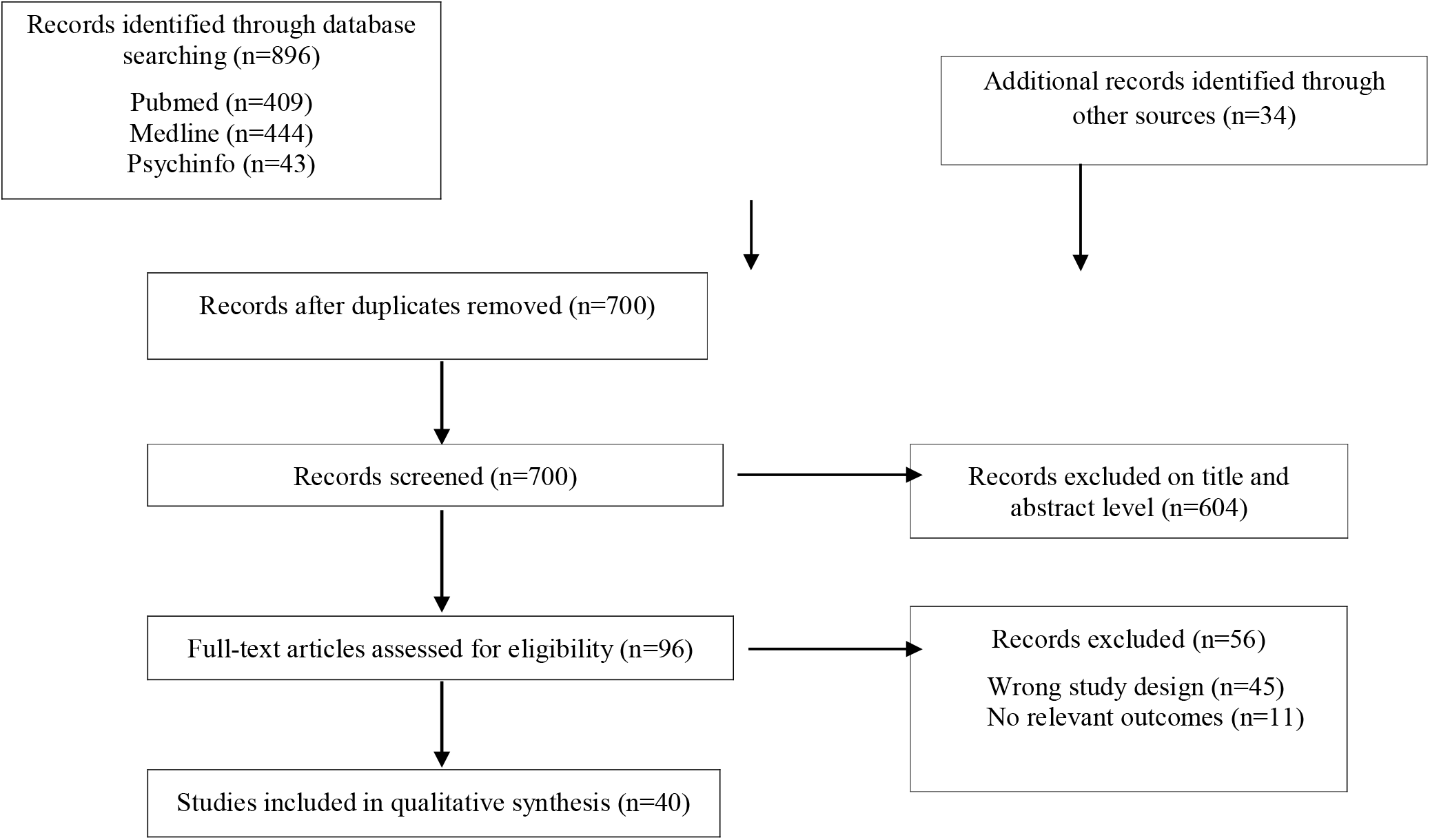
Study Selection.

Characteristics of the 40 studies included in the meta-synthesis are shown in Table 1.

Out of the 40 studies included in the meta-synthesis, 17 studies were based on participants in Asia, 12 in Africa, six in North America, three in Australasia and two in Europe. Fourteen studies looked at experiences related to SARS, 14 to Ebola, four to unspecified influenza pandemics, three to MERS, two at H1N1, two at COVID-19, one at Avian flu and one at swine flu. Most participants were described as healthcare workers, nurses or medical staff. All studies were published between 1999 and 2020. In most studies data was collected through individual interviews or focus groups, although one study was a personal reflective account and one paper a commentary citing interviews which had been conducted. A variety of analytic methods were used including thematic analysis, content analysis, framework analysis and phenomenological analysis, although many papers did not state the specific method used to analyse their data.

The quality of studies varied, although most were of moderate quality. The results of the quality assessment of included studies using CASP (2017) criteria is shown in table 2 (please see appendix for individual study quality ratings).

### Meta-Synthesis

The themes emerging from the meta-synthesis are shown in table 3.

**Table 3.** Overarching themes from meta-synthesis.

| **Themes (sub-themes)** |
| --- |
| 1. Physical health, safety and security<br>*1.1 Concerns for self*<br>*1.2 Concerns for others*<br>*1.3 Practical and environmental issues* |
| 2. Workload |
| 3. Stigma |
| 4. Ethical, moral and professional dilemmas |
| 5. Personal and professional growth |
| 6. Support to and from others<br>*6.1 Family and friends*<br>*6.2 Colleagues and peers*<br>*6.3 Organisations*<br>*Media and the public* |
| 7. Knowledge and information<br>*7.1 Communication*<br>*7.2 Training* |
| 8. Formal support |

### 1. Physical health, safety and security

Themes related to physical health, safety and security pervaded nearly all 40 papers.

#### 1.1 Concerns for self

The predominant concern across most staff groups was becoming infected with the virus themselves. Gershon et al. (2016) writing about healthcare volunteers’ experiences of treating Ebola in Emergency Treatment Units (ETUs) in West Africa describe:

> *Thoughts of getting infected were the uppermost concern for most, especially during the beginning of the deployment when they were still becoming acclimated to the ETU and whenever there was a breach in infection control protocol and practice. For some, fear was constant. One participant recalled constantly thinking, “Don’t let me get Ebola, don’t let me get Ebola*.*”*

Fears of contamination were exacerbated by experiences of inadequate PPE which was a recurrent theme across many papers, transcending different countries and pandemics. Shih et al. (2007) explored nurses’ experiences of treating SARS in Taiwan in the early 2000s and noted:

> *In this beginning stage, the factors contributing to the nurses’ fear about fatal infection by SARS were based on a lack of defensive protection against the disease*.

Thirteen years later, Yin & Zeng (2020) document nurses’ experiences of treating COVID-19 in China and quote one of their participants:

> *“I hope that personal protective equipment is available every day so that I don’t have to worry as much about myself or my colleagues getting infected*.*”*

#### 1.2 Concerns for others

A few studies provided exceptions where frontline workers reported less concern over their own immediate health, but nevertheless still expressed significant concerns for others. Workers were preoccupied about their families becoming ill and were particularly concerned that they themselves might transmit the illness to their loved ones. For example, talking about nurses’ experiences of SARS in Singapore, Koh et al. (2012) reported that:

> *Some participants were not concerned about themselves, rather they were concerned that they would, because of their exposure to infected patients, colleagues or visitors to the organization, inadvertently infect their family*.

Many made sacrifices and sought to protect their loved ones by staying away from them. For example, Yin and Zeng (2020) quote a nurse in China in the early stages of COVID-19:

> *“I stay at a hotel every day and am afraid of getting my family sick. I’m afraid to go home and haven’t seen my mom and dad for a long time*.*”*

Fellow healthcare workers falling ill with the virus was a significant cause for preoccupation and distress amongst participants.

> *All of the participants described being particularly vulnerable when caring for patients who were healthcare workers, whether doctors, nurses or support staff who had contracted the disease at work. That the patients were colleagues in a similar situation in life gave a more personally emotive dimension of the experience* (Erland et al., 2017).

#### 1.3 Practical and environmental issues

Healthcare workers were also impacted by practical and environmental issues in the settings in which they worked. Whilst, for the most part, healthcare workers’ fears were allayed by adequate PPE, it was also noted in several papers how the PPE caused discomfort and impacted on communication.

> *The equipment was described as cumbersome and hot, and staff reported finding it difficult to communicate with others who were wearing the PPE. Basic clinical procedures were deemed impossible by participants while wearing the recommended PPE* (Broom et al., 2017).

Some studies commented on the pay off between staff safety and patient care. For example, Moore et al. (2005) describing the treatment of SARS in Canada quote one participant:

> *“What we’ve been told is*…*that [in] triage, you change your goggles, gloves, mask and gown between every patient and its 100% not feasible. It can’t be done. Patients would be dying waiting at the triage desk*.*”*

Many studies also commented on the settings in which healthcare workers treated those affected by the pandemic being unfit for purpose and lacking in essential resources. Talking about nurses’ experiences of the Ebola crisis in West Africa, Gershon et al. (2016) state:

> *By any measure and at multiple levels, the early humanitarian response to the Ebola epidemic was extraordinarily challenging. Health care facilities and systems, already severely under-resourced in the affected areas, were strained to the limit*.

This was not unique to developing countries with similar challenges reported in Canada (Moore et al., 2005) and Australia (Broom et al., 2017).

### 2. Workload

Healthcare workers commonly reported elevated workloads, which impacted on their psychosocial wellbeing. They cited increased hours and weekend shifts, additional time taken to manage PPE and increased paperwork as frequent sources of stress. This was compounded by staff shortages (due to inadequate staffing or staff absences because of ill health or caring responsibilities) resulting in requirements for staff to work overtime. This led to the workers feeling fatigued and risking mistakes. A nurse from Toronto in the study by Moore et al. (2005) described her experiences:

> *“I work 12-hour shifts in emergency, rarely got a break, we were not permitted to have fluids at the desk. None. None in the care area. So we were going for five or six hours with nothing to drink. We were so exhausted. So at the end of your 12 hour shift by 6 or 7 hours you’re so exhausted that you’re crazy. That is now leading to sloppy practice*.*”*

However, because of staff shortages, some participants were noted to describe feeling guilty for taking time off to rest (Gershon et al., 2016; Rubin et al., 2016). Even when able to take a break, this was not always possible. Several studies described staff being unable to leave the hospital or hotel environment, feeling isolated outside of work or having little access to other activities (Rubin et al., 2016; Im et al., 2018).

Financial consequences of working, or not working, during the pandemic were also discussed. For some, sickness entitlements were considerably less than usual salaries and some staff were not paid at all if unable to work. This led to significant financial hardship and a motivation for some to work even when unwell. Some were offered a ‘risk allowance’ for the work that they undertook, which was a source of motivation for some, although professional duty of care usually outweighed financial incentives for most. Nevertheless, when financial remuneration was offered but delayed or not provided, workers felt *“abandoned”* and *“betrayed*.*”* Such betrayals of trust exacerbated pre-existing disaffection amongst healthcare workers.

> *“The SARS epidemic changed my view of nursing in Ontario. I finally realized that nurses were undervalued, underappreciated and undercompensated for the risks they take on daily to provide adequate healthcare to their clients*.*”* (Bergeron et al., 2006)

### 3. Stigma

Participants in many of the studies talked about experiencing stigma as a result of working on the pandemic. This was greatest in the earlier phases of the outbreaks or in contexts where less was understood about transmission of the virus.

> *In addition to their own fear of becoming infected with Ebola, the midwives also had to deal with the public fear of the contagious disease. Ebola was an unknown disease in Sierra Leone prior to the outbreak, and lack of knowledge resulted in rumours and misunderstandings among the general population* (Erland et al., 2017).

This stigma extended to the families of healthcare workers with some reporting their children being discriminated against (Kim, 2018). Other studies pointed to the hypocrisy that some healthcare workers experienced when they were publicly commended for their work but privately discriminated against (Im et al., 2018).

### 4. Ethical, moral and professional dilemmas

One of the greatest sources of tension was the competing obligation healthcare workers felt between providing good patient care and protecting their own physical safety. Strict infection control procedures meant that staff were not always able to intervene in the way they wanted, resulting in them feeling like they fell short of their usual standards of care.

> *“The biggest conflict within me, was the lack of ability…to put your hand on a shoulder…or hold their hand”* (Lamb et al., 2018).

Further constraints due to lacking resources exacerbated healthcare workers distress and led to a sense of futility. Comments such as *“I couldn’t do anything to help”* (Smith et al., 2017) and *“we could not do enough”* (Liu & Liehr, 2009) pervaded many narratives. Several studies discussed the undignified manner of patient deaths and healthcare workers’ lack of ability to provide access to adequate pain medication or give them any measure of comfort as a great source of distress.

Staff shortages and the associated lack of support meant workers were left to make difficult, often life and death, decisions on their own, which were noted to cause serious ethical dilemmas. Inequalities and decisions about who should get access to resources; beds, medication and vaccines also caused staff significant upset. The impact of these dilemmas appeared to continue after the crisis had passed.

> *Participants reported feelings of grief, mourning, sadness, depression, remorse, and regret upon their return. As one participant said, “Oh, we could have done much, much more*.*”* (Gershon et al., 2016)

Nevertheless, for the most part, workers felt inherently motivated to undertake this work and held a strong conviction that not doing so would be unethical. The idea of not treating patients affected by the virus was seen as *“cowardly”* (Lam & Hung, 2013) and *“morally unacceptable”* (Ives et al., 2009) and staff who avoided this work were viewed with scepticism.

### 5. Personal and Professional Growth

Concurrent to the pressures noted above, many healthcare workers described aspects of the work as enjoyable and rewarding and appeared to derive job satisfaction from work that they felt was *“important”* and *“meaningful”* (Bensimon et al., 2007).

These sentiments seemed more pronounced when workers saw patients improve and leave the ward, and over time as the number of infections and deaths declined. The gratitude of others; patients, their families and wider society was noted to increase their sense of fulfilment.

Even in some of the most challenging moments, many healthcare workers found meaning in their work, for example, Erland and Dahl (2017) describe midwives caring from pregnant women dying from Ebola in Sierra Leone who *“found it meaningful to be there and care for the women in their last moment of life*.*”*

Overcoming such immense challenges tested the participants and imbued some with a sense of greater professional confidence and competency. Several studies described staff gaining new knowledge and skills which they felt would equip them in their future work, especially if they ever worked in a pandemic situation again. Some also reported personal growth and developing confidence in their own resilience.

Nevertheless, deriving meaning and taking pride in their achievements did not render healthcare workers immune from the longer lasting impact of the work.

> *“I’ve just lost my way. When I got back, the problems were still there…Reforming a new life has been tough. I guess you could call it PTSD. I’m proud of what I did…. but in my personal life, I’ve paid a heavy price*.*”* (Gershon et al., 2016).

### 6. Support to and from others

Sources of support were discussed in many of the papers, although healthcare workers’ experiences demonstrated that many potential sources of support could also be additional sources of stress.

#### 6.1 Family and friends

Families and friends were important sources of support but could place pressure on the healthcare workers. Some tried to dissuade them from working on the pandemic, leading the workers, in some cases, to withhold from their families what they were doing.

For the most part, healthcare workers appreciated the opportunity to stay in touch with friends and family, usually over the phone. This was reported to bring them comfort as well as allay the worries of their loved ones. However, this could still place an emotional burden on the workers:

> *“Sometimes, I was too tired to talk over the phone but I still wanted to switch on the mobile because I was concerned about my family’s condition…I found I could not control my temper during that period. After two sentences of talk with my family, I felt short of breath and became very frustrated. I understood that my family would like to listen to my voice, but I just could not talk*.*”* (Mok et al., 2005).

The competing demands of managing work and family life during a pandemic was also a source of stress. As described by Bergeron et al. (2006) during SARS in Canada:

> *The juggling of work/family demands often had personal costs: “I rarely saw my husband and when I did I had little energy left for him. The strain almost cost me my marriage*.*”*

Reintegrating into normal family life after their work on the pandemic was over was also problematic for some. Several studies described workers missing colleagues and struggling to re-engage with previous work.

> *Others mentioned feeling isolated because the only people that they felt they could really talk to and who understood what they were feeling were the people who had deployed with them. “You breathe, you eat, you sleep it, for 24 hours of every day. It’s not like you can come back home and relax with your family. Your heart is just not into it*.*”* (Gershon et al., 2016).

#### 6.2 Colleagues and peers

For the most part, working with colleagues during the pandemic was noted to provide an important source of mutual support, opportunities to learn from each other and facilitate camaraderie.

Buddying systems, whereby more experienced staff supported newer staff, seemed appreciated, as were opportunities for informal group reflection. This seemed to enable staff groups to normalise difficult responses and provide appropriate reassurance. As described, in Lamb’s (2018) study of Ebola:

> *Participants described how they would simply sit down together at the end of a shift, share a cup of tea and discuss the events of the day: “quite a few of them [juniors] had never seen a dead body before, certainly never dealt with dying patients*... *but we would just sort of just sit down and chat about it and about how they were feeling…it was ok to feel upset, it’s just a perfectly normal reaction*.*”*

Some healthcare workers also spoke about the value of social media platforms for keeping in touch with colleagues, such as Whatsapp groups. Some did nevertheless lament the loss of previous opportunities to socialise with colleagues face to face and outside of work (Yin & Zeng, 2020).

Colleague and peer relationships could also be the source of some stress. Unfair distribution of work and the refusal of some colleagues to treat patients affected by the virus caused notable tensions.

> *‘‘There was real division created amongst staff. We’d all be working in the ICU and there was a long list of people who said they’re not going in because of so and so…And this created resentment, hostility because there were a core group of us who went in there more often than we would have had to otherwise had all of us been sharing that responsibility. We carried a burden that wasn’t equally shared*.*”* (Bergeron et al., 2006).

This was exacerbated by inequities in pay and conditions for what healthcare professionals perceived to be equivalent work with the same risks.

#### 6.3 Organisations

Healthcare workers valued support from their organisations but gave examples of not feeling adequately supported. Some workers reported feeling coerced into working with infected patients or in inappropriate conditions. Participants across the studies felt that their organisations had an institutional duty to provide staff with sufficient protection to work safely.

Workers reported feeling supported by their organisations when there was clear alignment and shared decision making between senior managers and frontline healthcare workers but less supported when staff safety was not a clear priority. Workers also valued their organisations supporting them to take time off from their roles.

Workers’ perceptions of their organisation’s preparedness varied with workers in several studies reporting a lack of established protocols. Staff in some studies commented on hoping that their organisations would learn from these experiences and be better prepared in the future.

Workers wanted their hard work and sacrifices to be recognised by their organisations, although the degree to which they expected to be additionally rewarded varied. Nevertheless, they expected a degree of support in return for the sacrifices they made that not all felt was met. For example, Guimard et al. (1999) commenting on a focus group discussion amongst nurses write:

> *It was revealed during discussions that most of the nurses who volunteered to care for Ebola patients were very disappointed about the recognition they received for their actions. Most of them felt abandoned by the managers of the hospital and felt they received insufficient financial and psychologic support during the epidemic*.

#### 6.4 Media and the public

The media’s portrayal of the pandemic had both positive and negative impacts. Some studies described the role of the media in perpetuating stigma. Al Knawy et al. (2019) writing about MERS in Saudi Arabia commented:

> *All participants referred to consistent and pervasive negative media commentary on the MERS-CoV outbreak…These negative commentaries were evident across local mass media (television, radio and newspapers) and social media - particularly Twitter. The negative media reporting was cited as negatively impacting staff morale and affecting workers socially and psychologically*.

Many healthcare workers felt that catastrophic portrayals of the pandemic on the news compounded families concerns. Such representations were also argued to be partly responsible for discouraging people to attend hospitals for other health concerns, to the detriment of public health and with financial repercussions for hospital departments.

The media, however, was often a source of information which healthcare workers found helpful, especially when they felt they were not party to information from their organisations. The media was also noted to be helpful in advocating for healthcare workers and mobilising resources, such as exerting pressure to provide more PPE.

The support of the wider public was considered vital and where the public did not comply with related directives this caused the healthcare workers anxiety and frustration. Bergeron et al. (2006) quoted one nurse from their study of SARS in Canada:

> *“My experience in the workplace regarding lack of compliance from clients in quarantine orders also makes me angry and afraid. I feel that even after all the work of ALL health care professionals, this issue may be impossible to be contained without support of the public*.*”*

Healthcare workers also sought recognition and validation from the public.

> *They wanted the public to know what they had been through and how they had put their own lives at risk to help protect others* (Gershon et al., 2016).

### 7. Knowledge and information

A pervasive narrative amongst the healthcare workers across all the pandemics was that of uncertainty, which precipitated and perpetuated fear and anxiety. Knowledge was key in decreasing uncertainty and many participants sought information, clarity and consensus with the purpose of achieving greater certainty.

#### 7.1 Communication

Communication was vital to the healthcare workers, however, not always experienced as helpful. Many reported inconsistent and ineffective messaging and a lack consensus between sources of information.

Ives et al. (2009) for example, report a lack of communication in their study of healthcare workers in the UK:

> *The majority of participants said they had been given neither information about pandemic influenza, nor been made aware of what would be expected of them during such a crisis, and this gave many the impression that their employing Trust did not care about them or take their needs seriously*.

Equally prevalent were comments about there being too much information. Rapidly changing and inconsistent information *“increased frustration and uncertainty”* (Chung et al., 2005). This resulted in *“confusion and lack of trust in the information received”* and subsequently *“dismissal of the information as clinicians were unable to assimilate the information in the limited time they had”* (Broom et al., 2017).

Communication was valued when it was centralised and co-ordinated and came from reliable authorities. Participants also valued leaders who were available and visible during the crisis.

How information was shared was also an important point, with healthcare workers pointing out that many staff did not have the time or access to be repeatedly checking emails. Clearly visible posters and information cascaded through team leaders at shift handovers were cited as helpful.

Healthcare workers also believed that communication is a two-way process and that their feedback and knowledge should be recognised and acted upon. They felt they should be consulted and involved in decision making and that their learning from doing this work on the frontline was vital for responding to the current as well as future pandemics.

#### 7.2 Training

Healthcare workers’ experiences of training were variable. For many, training imparted important information, allayed anxiety and facilitated greater confidence. Participants in the studies valued training in infection control procedures and safe use of PPE as well as more general training about the virus.

Participants in several studies, however, felt that they had not received adequate training. As one healthcare worker in Gershon et al.’s (2016) study of Ebola in West Africa described:

> *“They (the sponsoring agency) handed me a viral haemorrhagic fever guide. I read it on the plane, showed up, but I had no real idea of what I was doing*.*”*

Even though some participants described feeling unprepared, there was a sense in some studies of limited or superficial engagement with training. Training seemed better received when it was deemed as relevant, realistic and timely. Practical simulations increased workers’ confidence. Workers also highlighted the importance of learning through experience and commented on competence and confidence increasing over time.

### 8. Formal Support

The psychological impact on healthcare workers was acknowledged in many papers, however, few studies reported on workers’ experiences of any formal psychological interventions. The idea that mental health support would be available seemed to be important and helped to alleviate workers’ anxiety. For example, Yin and Zeng (2020) quoted one nurse in the early phase of the COVID-19 outbreak in China:

> *“I hope that the hospital sets up a psychological support task force to ease our tension and fears*.*”*

When psychological support services were mentioned, they seemed to be of most value when available on site, were flexible and informal, and were offered individually or in small group which fitted around the workers’ shifts. Workshops on coping and emotional support were also described positively in some studies. Some participants appreciated the availability of helplines, although others described these as too impersonal.

Even when formal support was available, some staff were ambivalent about engaging. Chen et al. (2020) in their commentary on medical staff in China in the early stages COVID-19 described:

> *The implementation of psychological intervention services encountered obstacles, as medical staff were reluctant to participate in the group or individual psychology interventions provided to them. Moreover, individual nurses showed excitability, irritability, unwillingness to rest, and signs of psychological distress, but refused any psychological help and stated that they did not have any problems*.

After the peak of the pandemic, the emotional impact of the work appeared to be acknowledged more. Workers in several papers were noted to report difficulties sleeping, experiencing invasive memories and ongoing hyper-arousal as well as struggling to adjust to being back at home and their normal work. Few described access to any kind of formal follow up, although when this was offered, this appeared to be appreciated. Even amongst those who described coping well and who did not want to engage with formal services, informal follow ups and check ins from their organisations and colleagues were valued.

> *After deployment, they stressed the need for mental health and psychosocial support, and they requested deeper knowledge about coping strategies. The respondents reported being focused on their duties and safety during deployment, and only allowing emotional reactions afterwards*. (von Strauss et al., 2017).

## Discussion

In this review we sought to better understand healthcare workers’ experiences of working on the frontline and their views about support during COVID-19 and comparable pandemics. We found 40 qualitative papers which met our inclusion criteria, and which covered a number of different pandemics over the past 20 years. Meta-synthesis revealed eight key themes which transcended temporal and geographical boundaries. Participants across all the studies were deeply concerned about their own and/or others’ physical safety. This was greatest in the early phases of pandemics and exacerbated by inadequate PPE, insufficient resources, and inconsistent information. Workers struggled with high workloads and long shifts and desired adequate rest and recovery. Many experienced stigma. Healthcare workers’ relationships with families, colleagues, organisations, the media and the wider public were complicated and nuanced and could be experienced concomitantly as sources of support but also sources of stress.

The results of this review show that the current experiences of frontline healthcare workers are not without precedent. The themes identified in this review from previous pandemics are remarkably resonant with what we are hearing about the impact of COVID-19 on healthcare workers across the world at the current time. This points to a potential mental health impact on staff that is comparable to that experienced in previous pandemics.

A recently published review and meta-analysis of the mental health impact of working on pandemics including SARS, MERS, Ebola and COVID-19 (Kisely et al., 2020) suggested that healthcare workers exposed to virus-related work are 1.7 times more likely to develop psychological distress and PTSD compared to non-exposed workers. Our review sheds light on potential risk factors and their mechanisms of effect including fear associated with threat to life, uncertainty due to inconsistent or rapidly changing information, and threat to integrity due to discrimination.

This review also shows that accessing social support, a previously well-established protective factor against mental health difficulties such as PTSD (Brewin et al., 2000), was complex. Workers often self-isolated to protect their loved ones, did not disclose details of their work to them, struggled to manage the competing demands of work and family life and felt like the people in their usual support systems could not relate to what they had been through. This compromised healthcare workers use of social support, which may potentially have a longer-term adverse impact on their mental wellbeing.

The ethical, moral and professional dilemmas that healthcare workers faced also increases their risk of ‘moral injury’. Moral injury has been defined as the psychological distress caused by actions, or inactions, which violate an individual’s moral code, or a sense of betrayal by others, and has been highlighted as a potentially significant concern for healthcare workers during COVID-19 (Greenberg & Tracy, 2000). The healthcare workers in this review were often unable to deliver the level of care they felt professional and morally obliged to provide and many felt betrayed by their colleagues, organisations and society. Moral injury is not in itself a mental health disorder but is a risk factor for further mental health problems and may be particularly pernicious in the context of a pandemic.

The results of this review also highlight potential protective factors. Healthcare workers valued clear, consistent, and compassionate communication. They engaged well with training when it was practical and specific. The felt valued by their organisations when they prioritised their safety and supported them with manageable workloads and time out from work. The wanted to be consulted and included in decision-making. Staff appreciated peer support and tended to seek emotional guidance from their colleagues. This draws attention to potential opportunities to further develop peer support systems and increase mental health awareness in the workplace. However, colleagues could also be a source of tension, so peer support interventions in this workforce warrant careful evaluation. We also need to carefully consider how peer-based interventions may work in such a crisis so as not to place an additional burden on the healthcare workers providing them when, by definition, they are going through the same stressors.

This review also demonstrates that psychological growth was possible. Most healthcare workers were inherently motivated to undertake this work due to a sense of professional duty. Many derived meaning and satisfaction from their work and reported learning and professional development. They also frequently reported strong team cohesiveness and camaraderie. As suggested by Gerada (2020) in a recent editorial comment, “some good must come out of COVID-19” and there is potential for greater recognition and appreciation of healthcare work.

Public attitudes and the media had both positive and negative impacts on the healthcare workers in these studies but have the potential to provide support and validation of their work. Nonetheless, gestures of solidarity, such as applause for healthcare workers which have been taking place around the world during COVID-19, have the potential to feel meaningless and hypocritical if support for healthcare workers from the public and government is not sustained after the pandemic.

One potential difference between previews pandemics and COVID-19 is that there is now greater acknowledgement of the mental health impact on healthcare workers and increasing recognition of the need to support their wellbeing (Billings et al, 2020). However, there is a yet little evidence about what is effective and what healthcare workers themselves want. The studies included in this review focused little on formal psychological interventions. It is therefore difficult to establish whether this was not of primary importance to the healthcare workers, whether they were not aware of sources of mental health support, or whether these interventions were simply not available to them. When mental health support was mentioned, participants tended to speak of it as desirable. However, some studies suggested a reluctance to engage. This is perhaps indicative of potentially enduring stigma amongst healthcare workers, exacerbated by militaristic metaphors and heroic narratives in the media, which make it harder for them to admit when they are struggling. This is an additional area that warrants exploration in order to better understand workers’ ambivalence and to ensure that they feel able to engage with appropriate mental health support when needed.

The results of this review also suggest that healthcare workers’ mental health needs change over time. In the early stages of the crises the workers prioritised more basic human needs, such as physical safety and rest. At the peak, workers seemed to focus on the work at hand and rely on colleagues for support. After the crisis had passed, there seemed to be greater recognition of the impact of working on the pandemic on mental health and an associated recognition of need for more support. At the current time, whilst attention is being paid to the impact on frontline healthcare workers’ mental health, there is a paucity of research into psychosocial interventions specifically for frontline healthcare workers and what works for whom and when.

We also know that the healthcare workforce was experiencing high levels of distress and disaffection prior to COVID-19 so we need to consider what should be set up as standard support for healthcare workers in the longer term. This will be particularly important if there are further waves of COVID-19, but also in the face of other, inevitable, future healthcare crises. We need to ensure that we maintain a psychologically healthy workforce, not just for the wellbeing of the workers, but also for the sustainability of healthcare services globally.

**Implications**

The findings of this review highlight a number of important implications which are relevant globally.

◼ Provision of adequate safety equipment is a priority to enable safe and effective working but also to mitigate negative mental health outcomes.
◼ Workloads need to be manageable, and sufficient periods of rest and recovery mandated to mitigate fatigue and burnout.
◼ Training should be relevant, practical, and timely. Learning on the job is valued alongside formal training.
◼ Communication needs to be clear and consistent and decision making shared. Leaders should be accessible and visible.
◼ Mechanisms to facilitate staff peer support should be put in place, including ringfenced time and mental health awareness training.
◼ Competing demands between work and family life should be acknowledged and staff supported in maintaining family roles as much as possible. Staff should be supported in taking time off from work.
◼ Anxiety, guilt, and moral injury may be mitigated by reducing lone working, encouraging buddying systems, facilitating ethical forums which allow workers to discuss difficult decisions and focusing on the meaningfulness of the work.
◼ Mental health follow-up will be imperative for the early detection and treatment of emerging mental health problems and to ensure staff feel supported by their organisations. Ongoing peer support is likely to be important.

## Strengths and Limitations

This review should be considered within the context of its strengths and limitations. This paper offers a rapid review of an important body of literature for the purposes of providing urgent feedback and guidance for those planning the support of frontline healthcare workers during the current COVID-19 crisis. We conducted our search across three databases and used two independent reviewers for searching, screening, data extraction and quality appraisal. We conducted a meta-synthesis for the reader to highlight overarching themes in relation to healthcare workers’ experiences of working on the frontline during a pandemic and their views about support.

There are a number of limitations inherent in the papers included. This review has highlighted a dearth of research exploring healthcare workers own views, needs and preferences. What research there is has focused predominantly on doctors and nurses with little or no identified research on other key frontline healthcare groups including physiotherapists, pharmacists, receptionists, porters or cleaners. Most of the studies were of only moderate-quality, therefore caution must be observed when considering the transferability of the findings.

There are also some limitations of the current review. Due to the rapidity required we did not pre-register a study protocol on Prospero. We also only searched a limited number of databases; therefore, some papers may have been missed, which may have provided more detail or contradicted the findings summarised here. While research from across the world was included in this review, we were only able to include studies published in English. This review may therefore be subject to some publication bias. Ongoing attention is warranted as papers reporting on this phenomenon may yet be published.

## Conclusions

This pandemic is not unprecedented. We have an opportunity to learn from the lessons of previous pandemics and provide better support for frontline healthcare workers. More high-quality qualitative research is urgently needed in order to better understand the experiences, needs and preferences of the healthcare workforce, particularly those frontline healthcare workers whose voices have not yet been adequately represented. We need to develop clinical guidance specific to supporting this workforce. This guidance should be developed in consultation and collaboration with the healthcare workers themselves. Interventions to prevent and treat mental health distress in healthcare workers need to be developed and their timing, effectiveness and acceptability carefully evaluated. We have an opportunity to mitigate the negative mental health impact of COVID-19 and support the longer-term wellbeing of the healthcare workforce across the world.

## Data Availability

All data included in this manuscript is secondary data already available in published form. Data derived from our own analysis is included in the manuscript and supplementary materials. Any further information required can be made available.

## Contributors

JB conceived of the idea for this review, conducted the meta-synthesis, wrote the draft of the manuscript and incorporated contributions from all the other authors. BCFC and VG completed the literature searches, screening, data extraction and quality appraisal under supervision of JB. TG and MB provided peer consultation on the design of the review. All contributed to validity checks on the meta-synthesis of the data. All authors commented on drafts of the paper and approved the final manuscript.

